# Evaluation of optical genome mapping in clinical genetic testing of facioscapulohumeral muscular dystrophy

**DOI:** 10.1101/2023.08.10.23292816

**Authors:** Anja Kovanda, Luca Lovrečić, Gorazd Rudolf, Ivana Babic Bozovic, Helena Jaklič, Lea Leonardis, Borut Peterlin

## Abstract

**Background:** Facioscapulohumeral muscular dystrophy (FSHD) is the third most common hereditary muscular dystrophy, caused by the contraction of the D4Z4 repeats on the permissive 4qA haplotype on chromosome 4, resulting in the faulty expression of the *DUX4* gene. Traditional diagnostics is based on Southern blot, a time- and effort-intensive method that can be affected by single nucleotide (SNV), and copy number variants (CNV), as well as by the similarity of the D4Z4 repeats located on chromosome 10. We aimed to evaluate optical genome mapping (OGM) as an alternative molecular diagnostic method for the detection of FSHD.

**Methods:** We first performed optical genome mapping with EnFocus™ FSHD analysis using DLE-1 labeling and the Saphyr instrument, for our patients with inconclusive diagnostic Southern blot results, negative FSHD2 results, and clinically evident FSHD. Secondly, we performed OGM in parallel with the classical Southern blot analysis for our prospectively collected new FSHD cases. Finally, we used panel exome sequencing to confirm FSHD2.

**Results:** By using OGM we could resolve two cases with diagnostically inconclusive Southern blot results as having shortened D4Z4 repeats on the permissive 4qA allele, consistent with their clinical presentation. Results of the prospectively collected patients tested in parallel using Southern blot and OGM showed full concordance, showing OGM to be a useful alternative to the classical Southern blot method for detecting FSHD1. For a patient showing clinical FSHD but no shortened D4Z4 repeat on the 4qA allele using OGM or Southern blot, a likely pathogenic variant in *SMCHD1* was detected using exome sequencing, confirming FSHD2. OGM and panel exome sequencing can be used consecutively to detect FSHD2.

## 1. Introduction

Facioscapulohumeral muscular dystrophy (FSHD) is an autosomal dominant disorder manifesting with atrophy and weakness of the facial, scapular, and foot dorsiflexor muscles **[1]**. FSHD is caused by the faulty post-embryonic expression of the *DUX4* gene from the permissive 4qA haplotypes. The pathogenic process is started through the inactivation of repression mediated by D4Z4 repeats located adjacent to the *DUX4* gene **[1]**. Two forms of the disease are known, FSHD1 and FSHD2, that differ in their molecular pathway but are indistinguishable clinically. In the case of FSHD1, which represents ∽95% of cases of FSHD, the de-repression of *DUX4* is mediated through the D4Z4 contraction on the permissive 4qA haplotype. The contraction is currently considered to be fully penetrant at ≤9 repeat units of the D4Z4 locus on the permissive haplotype, whereas 10-11 repeat units of the D4Z4 locus on the permissive haplotype are considered as reduced-penetrance alleles **[1]**. In the remainder, ∽5% of patients with FSHD2, the de-repression of *DUX4* is mediated by the hypomethylation of the D4Z4 repeats on the permissible 4qA haplotype, with some known causes of this hypomethylation being pathogenic variants in *SMCHD1* or *DNMT3B* genes **[1,2]**.

According to the current diagnostic guidelines from 2015 **[2]**, detection of the repeat contractions causing FSHD1 is routinely performed using restriction fragment digestion of peripheral blood lymphocyte DNA and Southern blot analysis with specific probes. FSHD2 can be diagnostically confirmed in individuals who possess at least one 4qA permissive allele containing unshortened, but hypomethylated D4Z4 repeats allowing for the faulty expression of the *DUX4* gene **[3]**. The D4Z4 hypomethylation is mediated by insufficient expression of *SMCHD1* or *DNMT3B* gene, and pathogenic single nucleotide variants (SNV), exon duplications or deletions in these genes can be detected using NGS, MLPA, and qPCR methods **[4,5]**.

Southern blot analysis is a time and labor-intensive method, that can be affected by SNV and CNVs adjacent to the restriction enzyme sites, as well as by the similarity of the D4Z4 repeats located on chromosome 10 **[6–8]**. Therefore, several novel methods, such as molecular combing, and optical genome mapping (OGM) have been evaluated regarding their detection of D4Z4 contraction, and recently been reported to be an accurate and reproducible diagnostic method for FSHD1 D4Z4 repeat contraction detection **[7–12]**. Implementation of novel methods into clinical diagnostics required repeatability, replicability, and reproducibility. The latter must be performed by independent validation. Therefore, our aim was to examine whether OGM can resolve our past cases with diagnostically inconclusive Southern blot results and to prospectively evaluate routine OGM performance in clinical diagnostics, by using a case-series comparison.

## 2. Materials and Methods

### Patients

The study was conducted according to the principles of the Declaration of Helsinki and was approved by the Institutional review board of the University Medical Center Ljubljana (grant 20210031). Informed consent for clinical testing, allowing the use of their anonymized samples for the development and improvement of clinical genetic testing, was obtained from the individuals included in the study during their clinical examination.

A total of 23 subjects were included in the study and tested using OGM, including three patients with FSHD phenotype and inconclusive Southern blot results and negative results for FSHD2, and 20 prospectively collected asymptomatic relatives or suspected FSHD patients. The prospectively collected patients consisted of routine cases referred for FSHD testing to the Clinical Institute for Genomic Medicine (CIGM), University Medical Center Ljubljana, Slovenia, and the asymptomatic family members of inconclusive Southern blot index cases. All samples were collected during 2021-2023. For families where the index case had an inconclusive Southern blot, only OGM was performed in the asymptomatic family members.

### Southern blot

Southern blot analyses were commercially performed by Leids Universitair Medisch Centrum, Leiden, The Netherlands (LUMC), as described on their website (https://www.lumc.nl/siteassets/over-het-lumc/afdelingen/klinische-genetica/bestanden/lab/specifieke-informatie-uitvoer-diagnostiek/moleculair-genetische-diagnostiek---spierdystrofieen.pdf).

Briefly, the DNA of the patient is digested using restriction enzymes EcoRI (GAATTC)/ BlnI (CCTAGG)/ ApoI (RAATTY), followed by Southern blotting and hybridization using locus-specific p13E-11 probes. When needed, the 4qA allele is measured using restriction fragment length digestion, Southern blot, and locus-specific 4qA and 4qB probes. Known limitations of the method include intactness of the p13E-11 probe DNA region on 4q35 loci and somatic mosaicism of the short 4q35 D4Z4 repeats. The reported results were obtained in the English language and give the repeat size of the 4q35 D4Z4 +/-1 repeat units (Table 1).

**Table 1.** Comparison of Southern blot and optical genome mapping for FSHD testing.

| Subject | Family | Indication | Southern blot result | OGM result |  |  |  |  |  |  |
| --- | --- | --- | --- | --- | --- | --- | --- | --- | --- | --- |
|  |  |  |  | 4qA |  | 4qB |  | 10qA |  | 10qB |
| P01* | F01 | FSHD | result inconclusive | 10 | 29 |  |  | 22 |  | 21 |
| P02* |  | FSHD | result inconclusive | 6 |  | 24 |  | 12 | 38 |  |
| P03* | F02 | FSHD | initial result inconclusive, repeated southern blot showed ~3 (+/-1) 4q35 D4Z4 repeat units | 3 |  | 30 |  | 10 | 20 |  |
| P04 | F03 | FSHD | ~8 (+/-1) 4q35 D4Z4 repeat units | 8 | 50 |  |  | 26 | 60 |  |
| P05 |  | FSHD | ~8 (+/-1) 4q35 D4Z4 repeat units | 7 |  | 36 |  | 2 | 12 |  |
| P06 | F03 | SF | ~8 (+/-1) 4q35 D4Z4 repeat units | 8 | 51 |  |  | 16 | 26 |  |
| P07 |  | A | familial D4Z4 contraction not detected | 56 |  | 32 |  | 15 | 36 |  |
| P08 |  | A | familial D4Z4 contraction not detected |  |  | 16 | 25 | 11 | 61 |  |
| P09 |  | A | familial D4Z4 contraction not detected |  |  | 15 | 50 | 11 |  | 27 |
| P10 | F01 | A | not performed | 10 |  | 32 |  | 35 |  | 21 |
| P11 | F01 | A | not performed | 29 |  | 32 |  | 34 |  | 21 |
| P12 | F02 | A | not performed | 42 |  | 29 |  | 6 | 20 |  |
| P13 | F03 | A | familial D4Z4 contraction not detected | 50 |  | 34 |  | 5 | 26 |  |
| P14 | F03 | A | familial D4Z4 contraction not detected | 29 | 51 |  |  | 26 |  | 31 |
| P15 |  | FSHD | not performed | 5 | 50 |  |  | 8 |  | 17 |
| P16 |  | FSHD | not performed | 9 |  | 53 |  | 26 | 27 |  |
| P17 |  | FSHD | No shortened 4q35 D4Z4 repeat detected | 15 |  | 23/24 |  | 21 | 23 |  |
| P18 |  | FSHD | not performed | 7 | 32 |  |  | 22 | 24 |  |
| P19 | F04 | A | not performed | 14 | 27 |  |  | 11 | 58 |  |
| P20 |  | FSHD | not performed | 6 |  | 15 |  | 6 | 22 |  |
| P21 | F04 | FSHD | not performed | 7 | 27 |  |  | 11 | 45 |  |
| P22 |  | FSHD | not performed | 4 |  | 35 |  | 6 | 12 |  |
| P23 |  | FSHD | not performed | 7 | 9 |  |  | 19 | 22 |  |

\*retrospectively included patients, FSHD - clinical FSHD, SF - suspected FSHD, A -
asymptomatic testing, relative(s) has/have FSHD, FXX- Family.

### Sequencing analyses

Patients with inconclusive Southern blot and/or suspected FSHD2 were evaluated for possible causative pathogenic variants in the *SMCHD1* gene by using sequence analysis of exons 1-48, commercially performed by Leids Universitair Medisch Centrum, Leiden, The Netherlands (LUMC), as described on their website (https://www.lumc.nl/siteassets/over-het-lumc/afdelingen/klinische-genetica/bestanden/lab/specifieke-informatie-uitvoer-diagnostiek/moleculair-genetische-diagnostiek---spierdystrofieen.pdf).

Additionally, exome sequencing was performed at the CIGM using a muscular dystrophies gene panel containing 211 genes, including *SMCHD1, DNMT3B*, and *LRIF1*, as previously described [13].

### Optical genome mapping

OGM was performed at the Clinical Institute of Genomic Medicine, University Medical Centre Ljubljana, Ljubljana, Slovenia. Briefly, for each sample, 1.5 million WBC from EDTA collected whole blood were used to purify ultra-high molecular weight (HMW) DNA using the SP Blood & Cell Culture DNA Isolation Kit following manufacturer instructions (Bionano genomics, San Diego USA).

The next day, HMW DNAs were labeled using the DLE-1 enzyme (CTTAAG) and DLS (Direct Label and Stain) DNA Labeling Kit (Bionano genomics). Between 4 and 12 ng/ul of HMW DNA solution per sample was loaded on a three-flow cell Saphyr chip and scanned on the Saphyr instrument (Bionano genomics). Saphyr chips were run to reach a minimum yield of 500 Gbp / sample with human GRCh38 as a reference. Molecule quality reports (MQR) were checked for adequate quality parameters and after this EnFocus FSHD specific pipeline was used to determine the length of the D4Z4 repeats and assign their haplotype on chr4 and chr10. The Saphyr instrument ran the ICS version 3. ICS 5.2.21307.1 for image processing. EnFocus™ FSHD Analysis was run on 1.7.1.1. version of Access and Solve3.7_03302022_283 versions was used on either the in-house server or on the compute on-demand option. The same versions of the software were used for all of the analyses performed.

## 3. Results

Thirteen patients with clinically evident FSHD, one clinically suspected FSHD patient, and nine asymptomatic relatives of FSHD patients were included in the study (**Table 1**). The asymptomatic relatives were from several families (**Table 1, Table 2)**. For families where the index cases had inconclusive Southern blot results, only OGM was performed in the subsequent asymptomatic family members. In total, Southern blot was performed for twelve individuals, and OGM was performed for all individuals included in the study. Full data from OGM DNA isolation, labelling, imaging, and assembly quality control steps is available in Supplement 1.

**Table 2.** Allele comparison in families with FSHD.

| Family | Subject | Indication | OGM result |  |  |  |  |  |  |
| --- | --- | --- | --- | --- | --- | --- | --- | --- | --- |
|  |  |  | 4qA |  | 4qB |  | 10qA |  | 10qB |
| F01 | P01 | FSHD | 10 | 29 |  |  | 22 |  | 21 |
|  | P10 | A | 10 |  | 32 |  | 35 |  | 21 |
|  | P11 | A |  | 29 | 32 |  | 34 |  | 21 |
| F02 | P03 | FSHD | 3 |  | 30 |  | 10 | 20 |  |
|  | P12 | A |  | 42 | 29 |  | 6 | 20 |  |
| F03 | P04 | FSHD | 8 | 50 |  |  | 26 | 60 |  |
|  | P06 | SF | 8 | 51 |  |  | 16 | 26 |  |
|  | P13 | A |  | 50 | 34 |  | 5 | 26 |  |
|  | P14 | A | 29 | 51 |  |  |  | 26 | 31 |
| F04 | P21 | FSHD | 7 | 27 |  |  | 11 | 45 |  |
|  | P19 | A | 14 | 27 |  |  | 11 | 58 |  |

In case of one the three patients with initially inconclusive Southern blot results for FSHD1 and negative results for FSHD2 testing, the repeated Southern blot performed at the LUMC correctly identified the shortened size of the repeat. In the remaining two patients with clinically apparent FSHD and inconclusive Southern blot results, this was either due to the lack of standard shortened 4q35 D4Z4 repeat indicating the possible presence of an alternative 4qA allele (P01), or the observation of a hybrid restriction enzyme band (P02), as per original Southern blot report. In both patients, FSHD2 was considered and excluded using testing for pathogenic variants in *SMCHD1*. Testing for other muscular dystrophies using exome sequencing was negative. In both cases, OGM showed a shortened D4Z4 repeat consistent with FSHD1 (**Fig 1**), in concordance with the observed FSHD phenotype in these individuals (**Table 1**). In both cases, the presence of additional SVs and CNVs were detected in the proximal chr4 region (Supplement 1), which was the most probable reason for the diagnostically inconclusive results of the Southern blot.

**Fig 1.**
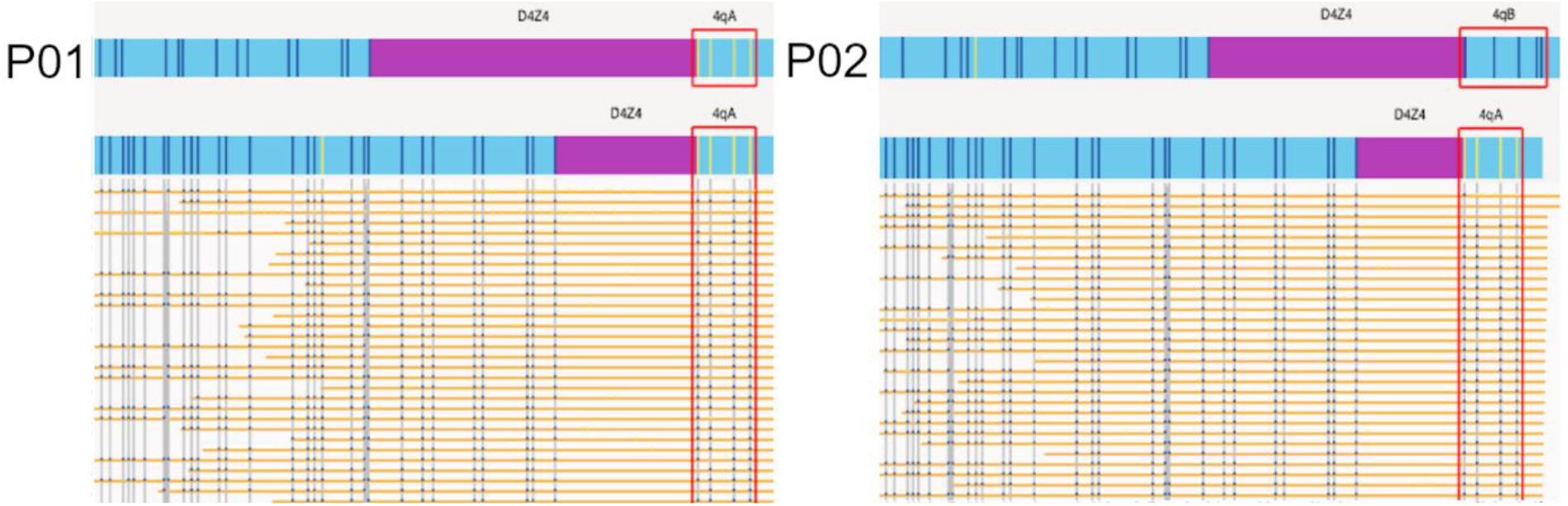
FSHD OGM maps of molecules at 4qA allele of P01 (left) and P02 (right). Individual molecules are shown for the 4q haplotype assemblies, showing support for the D4Z4 repeat size.

Of the prospectively collected patients, eight had clinically apparent FSHD and one had suspected FSHD, and by using OGM we showed eight cases had shortened D4Z4 repeats on the permissive 4qA allele, consistent with FSHD1, while in one patient (P17) no shortened D4Z4 allele was detected using neither OGM nor Southern blot on the susceptible 4qA allele. In this patient, exome sequencing performed at the CIGM and sequence analysis of exons 1-48 of *SMCHD1* performed commercially, concordantly detected the presence of frameshift variant c.2141dupC (p.(Ile715fs)) in the *SMCHD1* gene. The variant was classified according to the ACMG and AMP 2015 joint consensus recommendations [14] criteria PVS1 and PM2 as a likely pathogenic variant, and the diagnosis of FSHD2 was established in this patient, consistent with the clinical presentation.

In all prospectively collected samples, the results of OGM and Southern blot were completely concordant (**Table 1**), with both methods reporting a resolution of ±1 D4Z4 repeat. OGM FSHD results took an average time of one week, when included in a diagnostic lab workflow, and provided additional information on the length of D4Z4 repeats on the non-permissive alleles (4qB, 10qA, 10qB) (**Table 1**), which in our view is helpful information in subsequent testing of asymptomatic relatives.

In all asymptomatic family members, the results of OGM on all four alleles visible by OGM (4qA, 4qB, 10qA, 10qB) were consistent with allelic segregation and the alleles of the index case (**Table 2**). In family F01, one asymptomatic member was identified as having a shortened D4Z4 repeat in the reduced penetrance range.

The segregation of the alleles identified by OGM was also concordant in family F03, and in this family one suspected FSHD case was confirmed (**Table 2, Fig 2**).

**Fig 2.**
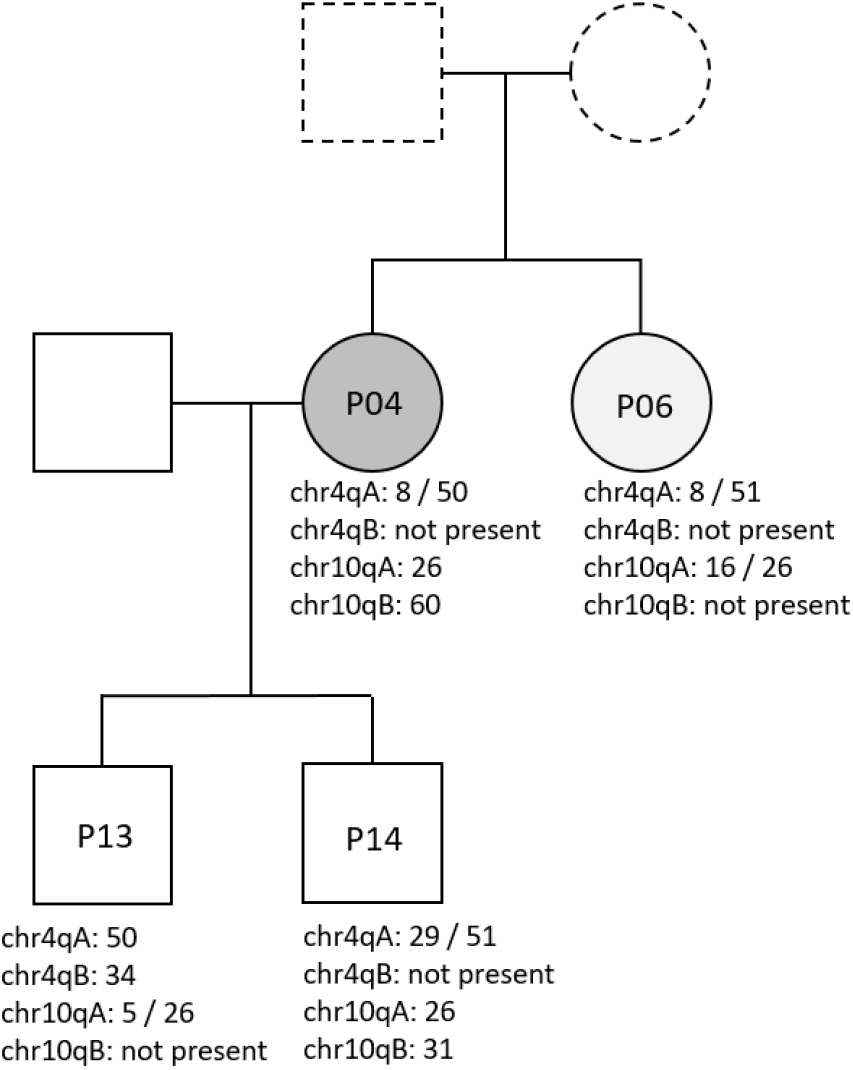
Pedigree showing segregation of chr4q and chr10q alleles in family 03. P04 - clinically apparent FSHD. P06 – suspected FSHD. P13 and P14 – asymptomatic testing. Parents of P04 and P06 were unavailable to perform testing.

FSHD - clinical FSHD, SF-suspected FSHD, A ^−^ asymptomatic testing, relative(s) has/have FSHD, FXX-Family.

## 4. Discussion and conclusion

According to the guidelines from 2015, Southern blot is defined as the routine genetic test in FSHD diagnostics **[2,15]**, despite recognized limitations such as sequence variation affecting probe-binding and restriction sites, somatic mosaicism, and hybrid alleles **[7,8,16]**. Other existing methods, such as molecular combing, PCR, and those examining hypo-methylation have so far not replaced this method, as they are effort-, time- and/or interpretation intensive, and may have limited cost-efficiency in the clinical settings with relatively low numbers of patients per year **[9,11,17–21]**. The advantage of OGM is that it can avoid some of these issues through simultaneous detection of all typical alleles on chr4 and chr10 in addition to providing information on additional FSHD-relevant regions such as CNVs in *SMCHD1* (which were not evaluated in this study)**[10,12]**. In the case of asymptomatic cases, accurate information on the presence of additional non-permissive 4q and 10q alleles provides, in our opinion, higher confidence to the FSHD-negative results.

In our study, OGM was concordant with clear Southern blot results in all cases. Additionally, we were able to resolve two additional cases of clinically evident FSHD with diagnostically inconclusive Southern blot results. In familial asymptomatic cases, OGM provided additional information on non-permissive alleles on chr4 and chr10 that segregated in an expected pattern, giving more confidence to the results of asymptomatic testing.

Based on our experience that OGM is a suitable alternative method to Southern blot for routine clinical use, we have incorporated OGM in the diagnostic approach to FSHD as the primary diagnostic test for FSHD1, followed by panel exome sequencing for genes involved in muscular dystrophies, including genes involved in FSHD2, as previously described **[13]**. In this way, we were able to successfully include OGM and exome sequencing into our diagnostic pipeline to detect a case of FSHD2.

As demonstrated by the accurate detection of the contracted repeats on the permissive 4qA allele using OGM in patients with inconclusive Southern blot results, OGM promises to improve the diagnostic outcome and accuracy in FSHD diagnostics. As with any new method, its limitations will become apparent with more data from its increased use for clinical genetic testing. Therefore, known and rare issues such as mosaicism, and translocations between chr4 and chr10, need to be kept in mind when interpreting results not consistent with the presenting phenotype **[22]**.

## Data Availability

The authors declare that the data supporting the findings of this study are available within the paper. Original data obtained and analyzed during the current study are available from the corresponding author B.P. on reasonable request.

